# Multi-channel intra-cortical micro-stimulation yields quick reaction times and evokes natural somatosensations in a human participant

**DOI:** 10.1101/2022.08.08.22278389

**Authors:** David A. Bjånes, Luke Bashford, Kelsie Pejsa, Brian Lee, Charles Y. Liu, Richard A. Andersen

## Abstract

Somatosensory brain-machine-interfaces (BMIs) can create naturalistic sensations by modulating activity of neural populations in the brain. By utilizing different spatial or temporal patterns of intra-cortical micro-stimulation (ICMS) in primary sensory cortex (S1), human patients suffering somatosensory loss can experience both cutaneous and proprioceptive sensory feedback. As evidenced by motor deficits in deafferented patients, rapid somatosensory feedback is critical for dexterous motor ability, in part because visual feedback is much slower than naturally occurring somatosensory input. However, somatosensory BMI studies typically report significantly longer cognitive processing latencies for cortical electrical stimulation than for naturally occurring somatosensations or visual sensations.

In this study, we show that multi-channel electrical stimulation patterns elicit naturalistic somatosensory percepts in a human tetraplegic participant. Crucially, somatosensations evoked by multi-channel ICMS are cognitively processed at comparable latencies to naturally evoked sensations and significantly faster than visual sensations, as measured via a simple reaction time test. Further investigation demonstrated multi-channel stimulation could significantly reduce minimum amplitude detection thresholds and such reductions in charge density resulted in more frequent “natural” sensation descriptors reported by the human participant. Multi-channel ICMS patterns also evoked percepts with highly stable somatotopic locations. While some single-channel ICMS patterns evoked sensations 20-80% of the time, most multi-channel patterns could evoke sensations with 100% repeatability, an important step in demonstrating BCI device reliability. These improvements are all significant advances towards state-of-the-art sensory BMIs. The addition of such low-latency artificial sensory feedback to motor BMIs is expected to improve movement accuracy and increase embodiment for human users.

## INTRODUCTION

Brain-machine-interfaces (BMIs) encompass a unique class of biomedical devices, enabling access to the user’s cortical neural activity through recording and stimulation [1]–[12]. These devices interface with the brain through a variety of modalities, including electrical [13], magnetic [14], ultrasound [15] or optical [16]. Electrical interfaces offer advantageous temporal and spatial properties for rapid, high-bit rate communication [17] or high degree-of-freedom (DOF) control of prosthetic devices [18].

BMIs users typically rely on visual cues to help guide their control of a computer cursor or prosthetic device [5], [10], [11]. With only visual feedback, BMI users experience similar obstacles to efficient, rapid, dexterous motor control as deafferented, sensory-impaired individuals [19]–[21]. Dexterous motor skills can be severely impaired by the presence of temporal delays in feedback, increasing task error and attenuating learning rates [22]–[24]. The long cognitive processing time required to process visual feedback for motor planning and execution likely contributes to the slow and clumsy control of BMI devices. Thus, restoration of sensory feedback with rapid cognitive processing latencies and naturalistic perception is a high priority for BMI development [25], [26].

Intra-cortical micro-stimulation (ICMS) has been shown to evoke sensory percepts via electrical stimulation patterns in both animal [27]–[30], [31]–[34] and human studies [3], [12], [35]. In animal studies, discrimination tasks have demonstrated that ICMS patterns can evoke a somatosensation indiscriminable from a mechanically produced sensation. In a seminal study, Romo and colleagues created a flutter sensation on the finger-tip of a non-human primate (NPH) by modulating the frequency of electrical stimulation [36]. Building on this work, researchers created an electrical stimulus which elicited the mechanical sensation of pressure in the finger-tip. By mapping the receptive field of primary sensory cortex (area 3a) of an NPH, they modulated the amplitude of the electrical stimulation pattern to correspond to the intensity of pressure [37]. In clinical studies, human participants can simply verbally report a qualitative description of somatosensations evoked by ICMS patterns [3], [12], [35]. The wide variety of cutaneous and proprioceptive somatosensations reported by participants in these studies underscores the importance of involving human participants in further BMI development.

Feasibility studies have demonstrated the successful integration of both sensory and motor control components in a single BMI device [30], [38]–[41]. However, typical single-channel ICMS patterns require significantly longer cognitive processing time compared to naturally occurring sensations or visual cues [42]–[45]. One study has demonstrated that even with such limitations; BMI control could be improved for a ballistic motor task [41]. For fine motor tasks however, such as dexterous object manipulation, single-channel ICMS may not be able evoke sensations fast enough to enable improvements to BMI control. If the cognitive processing latency of ICMS stimuli is longer compared to visual feedback, it is unlikely to be widely utilized by BMI users. Reaction times between multi-channel ICMS and naturally occurring stimuli have been comparable in a non-human primate study [45]. However, benchmark performance measures for reaction times to sensations evoked by ICMS have varied in other studies [42], [45], [46].

Elucidating fast reaction times in human participants is of high significance for the development of sensory BMIs. To this end, we sought to quantify differences between single- and multi-channel ICMS patterns delivered to primary sensory cortex (S1) of a human participant with tetraplegia. Electrical stimulation patterns were delivered on groups of electrodes across two chronically implanted microelectrode arrays. Using classical psychometric methods, we measured the participant’s reaction times from ICMS evoked sensations compared to visual and vibrotactile stimuli. Since single- and multi-channel stimulation patterns might evoke different sensory experiences, we stimulated channels across both arrays and quantified differences between verbally reported evoked somatic sensations. To elucidate the role of charge density when stimulating across multiple electrodes, we examined differences in minimum detection thresholds, the variability of somatotopic location for a given stimulation pattern, and modulation of the “naturalness” of the verbally described percept.

## RESULTS

A right-handed human participant with a C5-level spinal cord injury was implanted with two microelectrode arrays (Blackrock Microsystems, UT, USA), each containing 48 electrodes arranged in an 7×7 grid and coated in sputtered iridium oxide film (SIROF) [47], [48]. After implantation in left primary sensory cortex (S1), maps of the receptive field of each array showed coverage of upper and lower regions of the contralateral arm and several locations on the palm [3]. The current study examines somatosensations evoked via both single- and multi-channel intra-cortical micro-stimulation (ICMS), approximately 3 years post-implantation.

### Reaction time task

To measure the cognitive processing latencies of several sensory feedback modalities, we compared the simple reaction times (RTs) of cutaneous sensations evoked via ICMS, visual stimuli and naturally occurring cutaneous stimuli (Figure 1A). We used three different cortical stimulation patterns to capture RTs from single- and multi-channel ICMS patterns. One stimulus type was presented each trial and stimulus types were pseudo-randomly interleaved. The participant was cued at the start of each trial to attend to the particular stimulus type. Since voluntary attention can modulate reaction times [49], a fixation circle was presented along with a target circle, prior to stimulus presentation, in line with previous studies [45]. Since the tetraplegic participant had limited volitional control of their arms and was unable to reliably press a button, they were instructed to saccade from the fixation circle to the target circle when the stimulus presentation was perceived.

**Figure 1:**
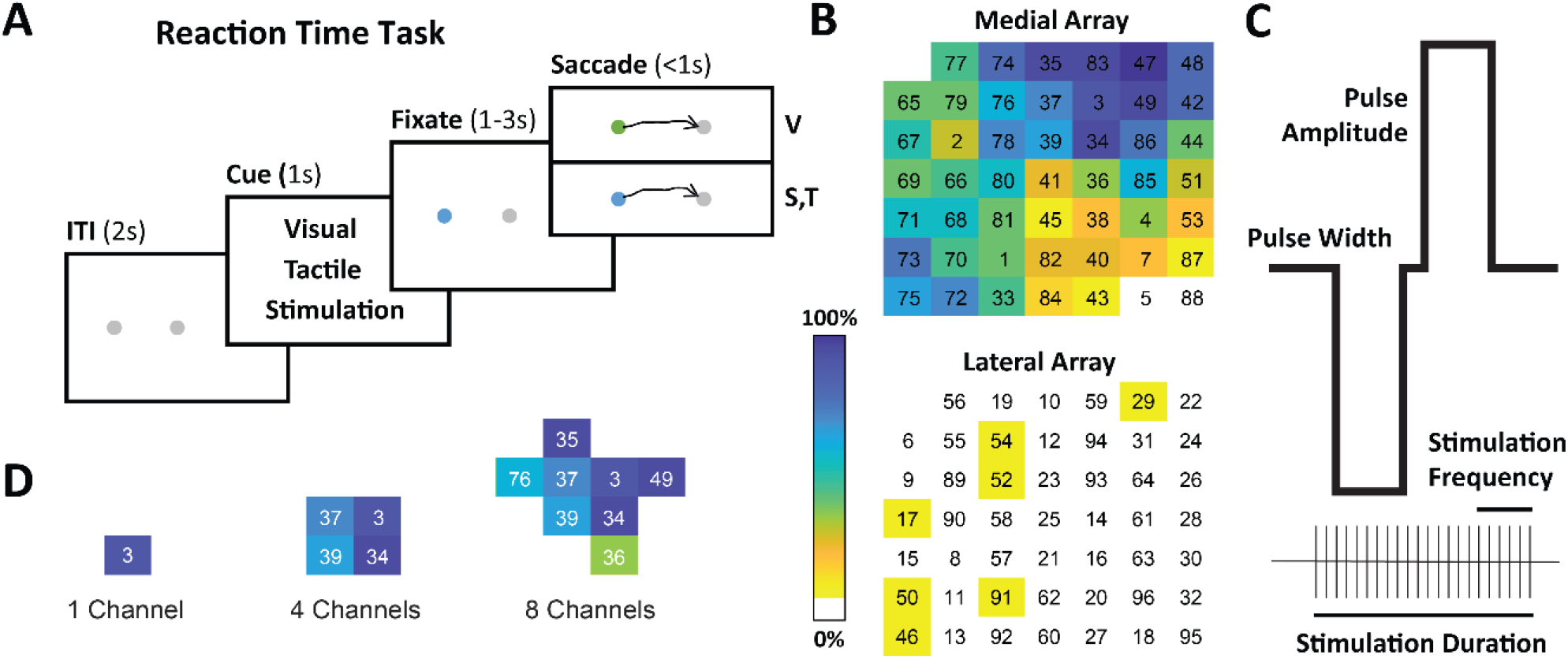
Reaction Time Task. (A) The task had four phases: an inter-trial interval, cue, fixation, and stimulus phase. The subject was cued to attend to one of three stimuli conditions. During the fixation phase, both the fixation target and the saccade target were present while the participant was instructed to fixate on the fixation target. After a variable fixation period, the participant saccaded towards the saccade target, once they detected the onset of the stimulus. An eyetracker was used to capture trajectory information and measure the participant’s reaction time. Three types of stimuli were used: visual (a change in the color of the fixation point), ICMS and vibrotactile stimulation. Each type of stimulus was pseudo-randomly interleaved trial per trial. (B) A spatial heatmap of both micro-electrode arrays implanted in S1. The channel numbers are displayed in the 7×7 grid of each array. The color index (blue to yellow) displays the probability of evoking a somatosensory response from various single-channel stimulation paradigms. (C) The ICMS stimulation pattern was designed with five main variables: pulse-amplitude, pulse-width, stimulation frequency, stimulation duration, and number of electrodes. (D) Groups of channels used in the ICMS reaction time stimulation condition. RTs to the single channel group were compared the two multi-channel groups in Figure 2. These neighboring channel groups were chosen based on channels with a high probably of evoking a sensation from the map in part B.

To provide a fair comparison between the RTs from each type of stimulus, parameters were chosen to maximize the experienced intensity, since stimulus intensity has been shown to modulate RT [50]. The three ICMS stimulation patterns evoked highly-stereotyped, cutaneous sensations, across all reported qualities: intensity, description and somatotopic location. Each pattern delivered 100µA, 300Hz, 200ms pulse trains to electrodes in one of three groups (Figure 1D). This corresponded to a total charge delivery of 1.2, 4.8 or 8.6µC (the eight channel group delivered only 90µA to each electrode due to charge safety constraints).

To mechanically produce a naturally occurring stimulus, we used a vibrotactile tactor (C-3 Tactor – Engineering Acoustics, Inc) to create a stereotyped cutaneous sensation. We chose a vibration frequency of 250Hz, known to evoke a strong response in Pacinian corpuscles and commonly used in across psychometric literature [51], [52]. The highest possible sinusoidal amplitude of the device was used without causing perceivable auditory artifacts. The device was capable of inducing a maximum possible deflection of 0.55mm, peak to peak. Two locations were tested independently. One on the upper bicep was closest to the ICMS evoked locations where the participant still had natural sensation (see methods) and the second was on the cheek.

For both ICMS and vibrotactile stimuli presentation, both the fixation and saccade targets did not change in visual appearance during the trial. For the visual cue presentation, the fixation circle changed from blue to green. This significant change in color has been shown to be highly discriminable in psychometric tests in healthy human participants [53].

### Multi-channel ICMS produced the fastest reaction times

The eight-channel ICMS pattern had the fastest measured RT range of any stimulus type (lower c.i. 167ms, Figure 2). Reaction times for both four- and eight-channel ICMS (median 187ms, 195ms) were significantly faster than single-channel RTs (median: 270ms) and significantly faster than the visual stimulus RTs (median: 309ms) (p < 0.001, Wilcoxon rank sum test with Bonferroni correction). The four-channel ICMS RTs (median: 187ms) was significantly than the RTs from naturally evoked vibrotactile sensations (median: 211ms) on a comparable location of the arm (p < 0.05, Wilcoxon rank sum test with Bonferroni correction). This ∼20ms difference roughly corresponded to an estimated neural conduction delay from the arm to cortex [45].

**Figure 2:**
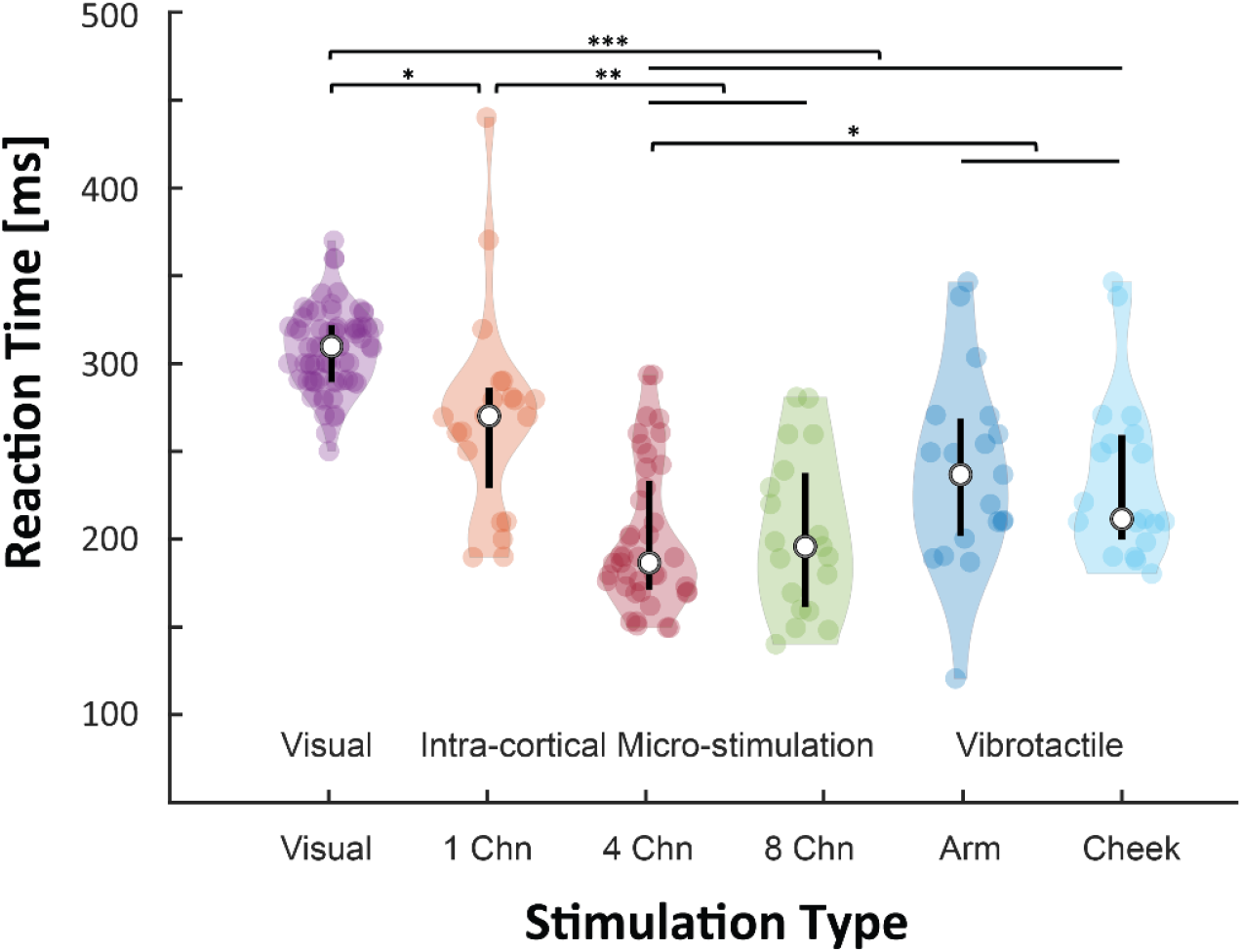
Reaction times and stereotyped ICMS evoked sensations. We directly compared reaction times to three types of stimulus presentation: a visual cue, a cutaneous vibrotactile sensation and cutaneous sensations evoked by ICMS. Parameters for each stimulus type were chosen to maximize intensity and thus produce the fastest reaction time, within the constraints of the task design. Single- and multi-channel ICMS patterns evoked highly stereotyped sensations. Neighboring channels with a high probability of evoking a sensation were chosen for each of the groups (Figure 1D). ICMS patterns evoked sensations in the inner elbow and lower foream. Vibrotactile stimuli were presented via a tactor placed on the arm and cheek. The vibrotactor arm location (upper bicep) was selected as closest to the ICMS evoked locations where the participant still experienced natural sensation (see methods). The visual cue was presented via a change in color. Reaction times were measured (median, 95% confidence interval) as follows: visual cue (309ms, c.i. 300-318mms), single-channel (262ms, c.i. 207-270ms), four-channel (187ms, c.i. 178-202ms), eight-channel (195ms, c.i. 167-231ms), arm (236ms, c.i. 207-262ms) and cheek (211ms, c.i. 206-255ms). Individual dots show each trial with the median and quartiles (25%/75%) alongside. Twenty trials for each type were measured except for visual and four-channel stimulation which had 70 and 40 trials respectively. Significance testing was performed with a Wilcoxon rank sum test while correcting for multiple comparisons with the Bonferroni method (* p < 0.05, ** p < 0.01, *** p < 0.001).

Measured RTs from the cheek (median: 211ms) were faster than RTs from the arm (median: 236ms). Due to the number of neural connections between mechanoreceptors in the periphery and primary sensory cortex and accounting for axonal conduction velocities, this was a typical result [54]. Both vibrotactile RTs were significantly faster than those from the visual cue (median: 309ms), as expected.

### Percepts evoked via ICMS during the reaction time task

At the end of each ICMS trial, the participant verbally reported each evoked sensation’s intensity on a 1-10 scale (10 being the highest), somatotopic location and description. Single-channel stimulation yielded an average intensity of 1.85±0.96 (mean ± standard deviation), while four-channel and eight-channel were each significantly higher, averaging 6.17±1.75 and 5.70±1.30 (p<0.0001, ANOVAN test, Bonferroni correction).

Single- and multi-channel ICMS patterns used for the reaction time task evoked highly stereotyped sensations. Single- and four-channel ICMS patterns elicited sensations on same somatotopic location on the arm (interior of the right elbow) with a 70% and 85% rate, respectively (Figure 2). Eight-channel stimulation elicited sensations on the right posterior forearm at a 100% rate.

Single-channel ICMS evoked a cutaneous sensation of “squeeze”, “touch”, “grab” or “pinch” with a greater than 90% rate. Four-channel ICMS evoked a cutaneous sensation of “squeeze” or “pinch” with a greater than 93% rate. Eight-channel ICMS evoked a reported sensation of “squeeze” with a 95% rate.

In the following sections we quantified the reported descriptions, somatotopic locations, and perceptual sensitivity of percepts evoked by multi-channel compared to single-channel stimulation.

### Verbal report task

Previous human studies reporting evoked ICMS sensory percepts, somatotopic receptive fields were mapped by stimulation patterns delivered to each electrode individually [3], [12], [55]. Due to constraints on the amount of electrical charge delivered safety through one electrode [56]– [58], a fundamental limit exists on the volume of neural tissue that can be stimulated. Possibly, this charge limit could exclude the parameters of evocable sensations or somatotopic locations only accessible by stimulation of a larger volume of tissue. To test this hypothesis, we stimulated both single- and multi-channel ICMS patterns on electrodes across both implanted arrays and compared the participant’s verbal report of each evoked sensation’s somatotopic location (Figure 3A).

**Figure 3:**
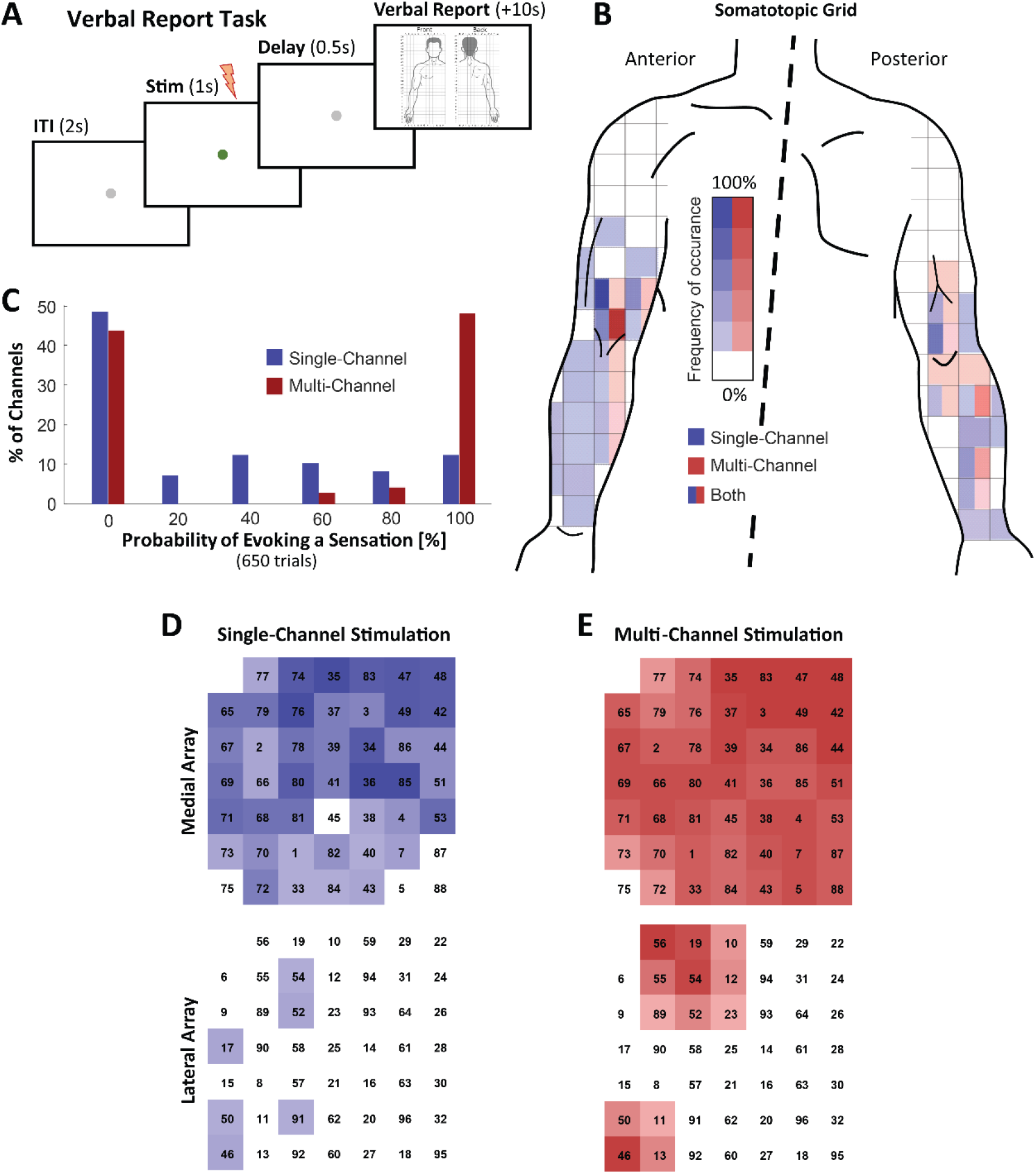
Array-wide sensitivity to single-or multi-channel stimulation. To directly compare receptive fields and somatotopic localization, both S1 arrays were stimulated with a single-or four-channel group (arranged in square of neighboring channels). If a channel (or group of channels) evoked a sensation, the participant verbally reported the location on a somatotopic body map. **(A)** The verbal report task consisted of four phases: an inter-trial interval (ITI), a stimulation pattern followed by brief delay and then a verbal report from the participant about the evoked sensation. **(B)** Probability of a body location reported from ICMS are shown in blue (single-channel) and red (four-channel) for each square grid location. If a square is a solid color, only one type of stimulation pattern activated this somatotopic region. If both colors are shown, both types evoked a sensation in this location. Darker colors indicate a higher probability of a sensation occurring at that spot. **(C)** Histogram of sensation likelihood. Blue bars (single-channel) and red bars (four-channel) indicate the percentage of channels for each stimulation type which had high and low probability of evoking a sensation. Multi-channel stimulation had a higher percentage of channels (48%) which responded with 100% likelihood than single-channel (13%). **(D)** Probability of evoking a sensation by stimulating each electrode individually. Darker colors indicate a higher probability of evoking a sensation. Five trials were measured on each electrode across both arrays. **(E)** Same analysis as part C for multi-channel stimulation patterns. Five trials for each square group of channels were measured.

### Across both arrays, multi-channel ICMS more reliably evoked a sensation

We mapped the receptive fields across all electrodes on our implanted arrays using our verbal report task (Figure 3A). We sequentially stimulated each single-or four-channel group and instructed the participant to indicate the somatotopic location for each evoked sensation on a body map (Figure 3B). Both the single- and four-channel stimulation patterns delivered a 100µA pulse-amplitude, 300Hz stimulation frequency for a one second duration on each electrode stimulated. Each four-channel stimulation pattern was delivered on four neighboring electrodes arranged in a square.

Across both micro-electrode arrays implanted in S1, we saw a substantial increase in the probability of evoking a sensation when using four-channel stimulation patterns (Figure 3C). While single-channel stimulation elicited a wider range of somatotopic areas, novel locations were reported during the four-channel stimulation that were not reported during the single-channel stimulation (complete red squares in Figure 3B). These data suggest single-channel stimulation alone is not sufficient to activate all possible somatotopic locations available with an implanted array. Heatmaps of both arrays shows definitive spatial organization of channels likely to evoke sensations (Figure 3D,E). We observed a higher concentration of activatable channels centered in the upper right of the medial array and sparse locations on the lateral array.

### Spatial patterns of ICMS analyze role of charge in evoked somatosensations

Previous studies have suggested electrical charge as a powerful modulator of somatosensory sensations in single-channel ICMS [28], [59], [60]. Specific spatial patterns of charge, delivered over multiple ICMS channels, may be an important parameter for targeting specific percepts or somatotopic locations [34][39]. While previously unexplored in human participants, we tested this hypothesis by directly comparing detection thresholds of various single- and multi-channel ICMS patterns using the verbal report task (Figure 3A).

We selected a group of 5 highly reliable electrodes (from the heatmap in Figure 1B) to quantify the effects of spatial patterning. Within this set of five electrodes, we selected 10 sets of paired electrodes, 5 sets of 4 electrodes and 1 set of five electrodes (Figure 4A).

**Figure 4:**
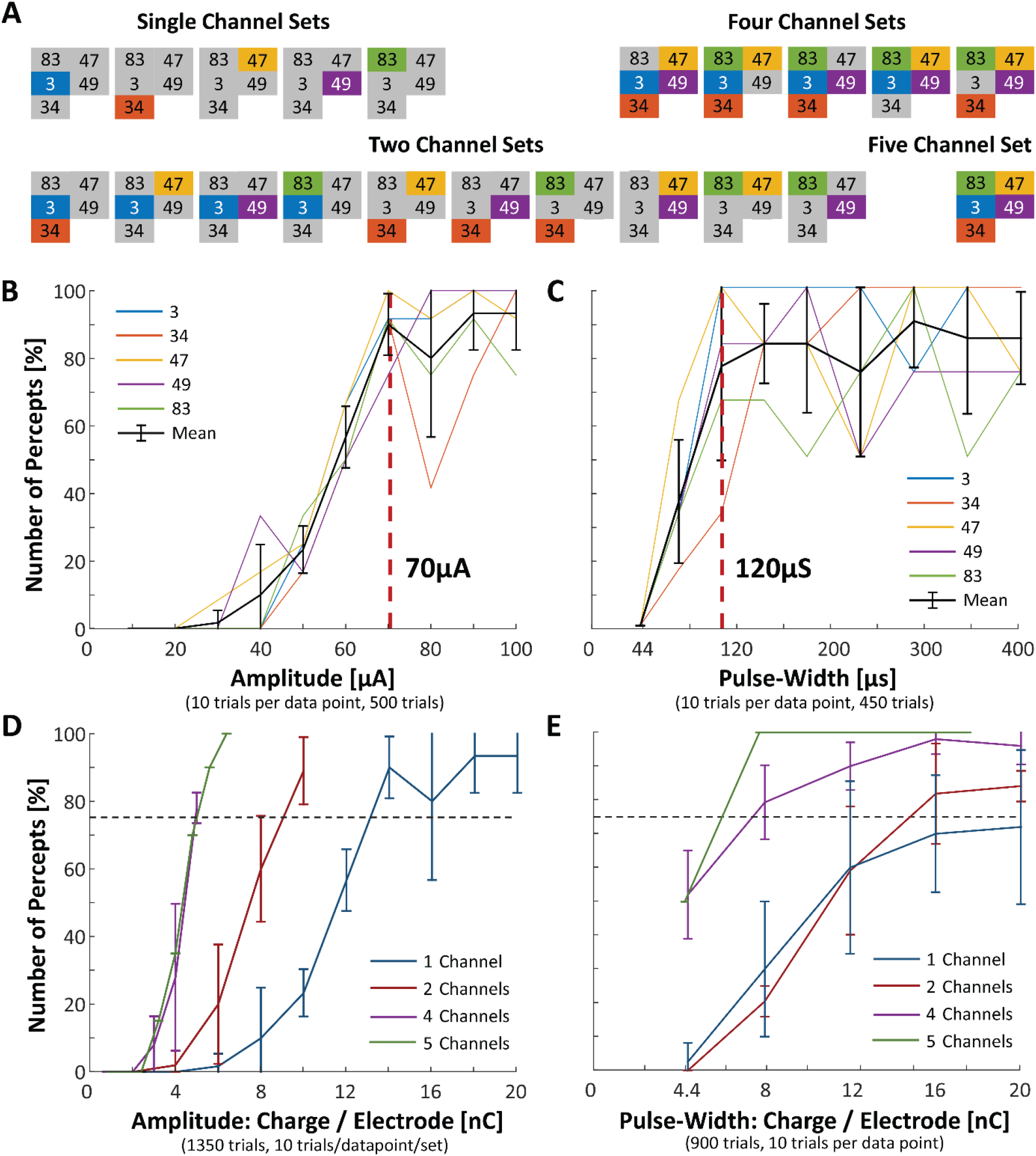
Probability of sensation for single- and multi-channel stimulation paradigms. The verbal report task was used to collect two datasets. Each trial contained a single stimulation pattern lasting for 1 second. The participant verbally responded if they felt a sensation. **(A)** Five highly responsive neighboring electrodes were selected (Figure 1C). Single and multi-channel groups were chosen for analysis in Parts D,E. Panel E used only the first four sets of the two channel pairings due to time constraints on data collection. **(B)** Psychometric threshold detection curves for pulse-amplitude modulated, single-channel ICMS patterns. The average threshold value across these electrodes was 70µA, corresponding to a total charge delivery of 14nC over one second of stimulation at 300Hz. Each channel was color coded and the average and standard deviation was shown in black. Pulse-width was fixed at 200µs for all stimulation patterns. **(C)** Psychometric threshold detection curves for pulse-width modulated, single-channel ICMS patterns. Pulse-amplitude was fixed at 100µA and sensations were reliably evoked with pulse-widths of 120µs or greater. **(D)** Psychometric threshold detection curves for pulse-amplitude modulated, multi-channel ICMS patterns. Each line shows the average response (mean and standard deviation) of all channel combinations in each respective group. Ten trials were averaged for each combination in each group. Trials from the single-channel group totaled 500, the two-channel group totaled 500 trials, the four-channel group totaled 300 trials, and the five-channel group totaled 50 trials. The y-axis shows the percentage of reported percepts for each condition/data point. The x-axis shows the amount of charge delivered per each channel. See methods for detailed explanation of charge calculation. **(E)** Psychometric threshold detection curves for pulse-width modulated, multi-channel ICMS patterns. Pulse-amplitude was fixed at 100µA. Trials from the single-channel group totaled 250, the two-channel group totaled 400 trials, the four-channel group totaled 200 trials, and the five-channel group totaled 50 trials. Since only one set of five electrodes was tested, no error bars are shown. Similarly, to Part D, the required charge per electrode to reliably evoke a sensation was significant decreased for multi-channel ICMS patterns.

To elucidate the role of charge density, we fixed the total amount of charge delivered across all channels, either concentrating it all on a single electrode (Figure 4B,C) or dividing the charge equally across two, four or five channels (Figure 4D,E). From these two datasets, we analyzed the likelihood of evoking a sensation (Figure 4), the reported descriptions of evoked sensations (Figure 5) and their somatotopic locations (Figure 6).

**Figure 5:**
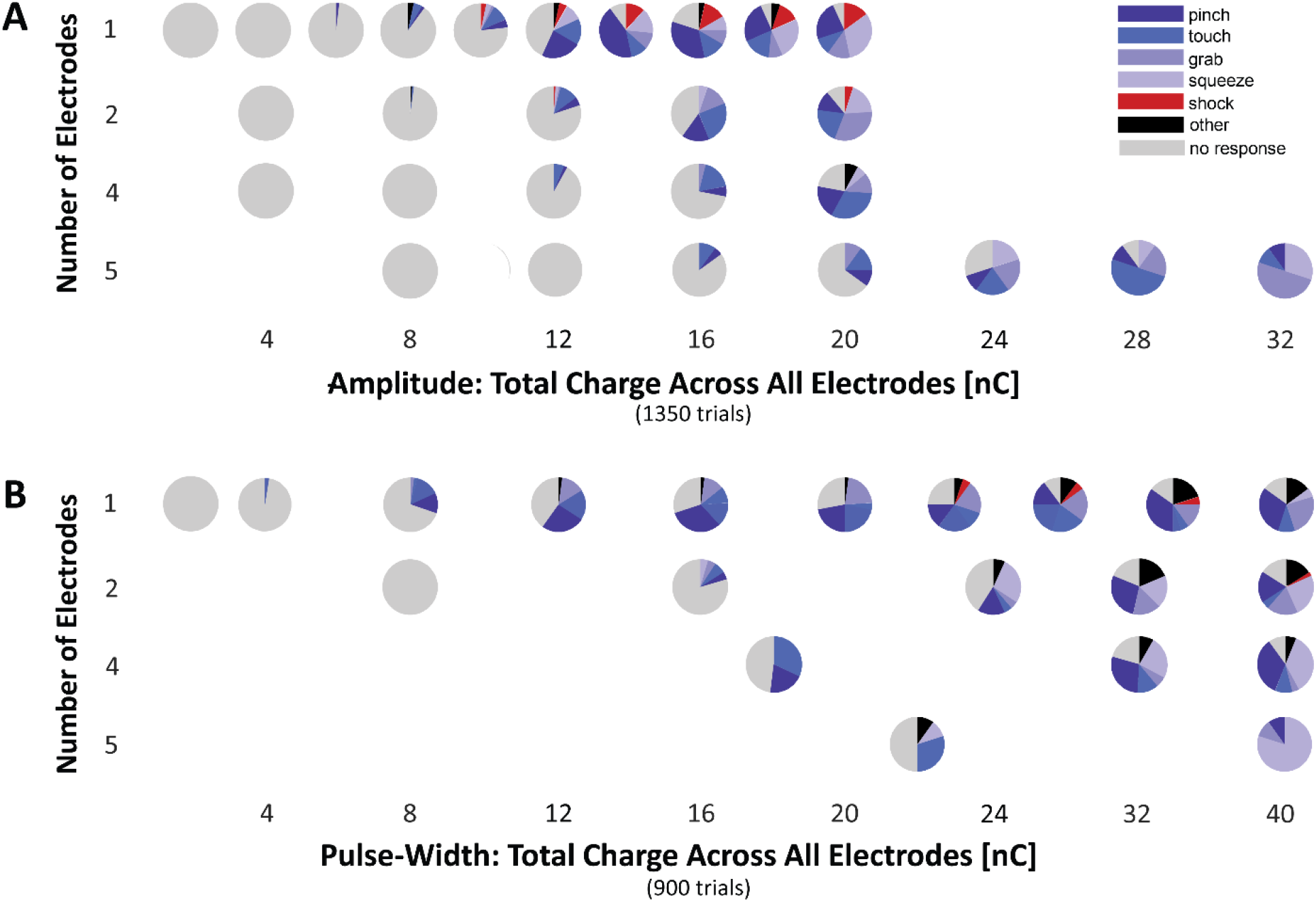
Reported descriptions of evoked sensations during single- and multi-channel stimulation. “Naturalistic” descriptors of evoked sensations were overwhelming reported for both single-channel and multi-channel ICMS. The frequency of “natural” sensations increased inversely proportionally to charge per electrode for pulse amplitude. As total charge increased on single-channel ICMS patterns, more “non-natural” sensations were reported. **(A)** Each pie chart represents the percentage of reported descriptions to single- and multi-channel ICMS patterns. Data were averaged over all trials for each data point shown in Figure 4C. “Shock” was the only “non-natural” descriptor reported, indicated in red. “Naturalistic” sensations are shown in shades of blue. **(B)** Identical analysis as part A, for the data collected in Figure 4D.

**Figure 6:**
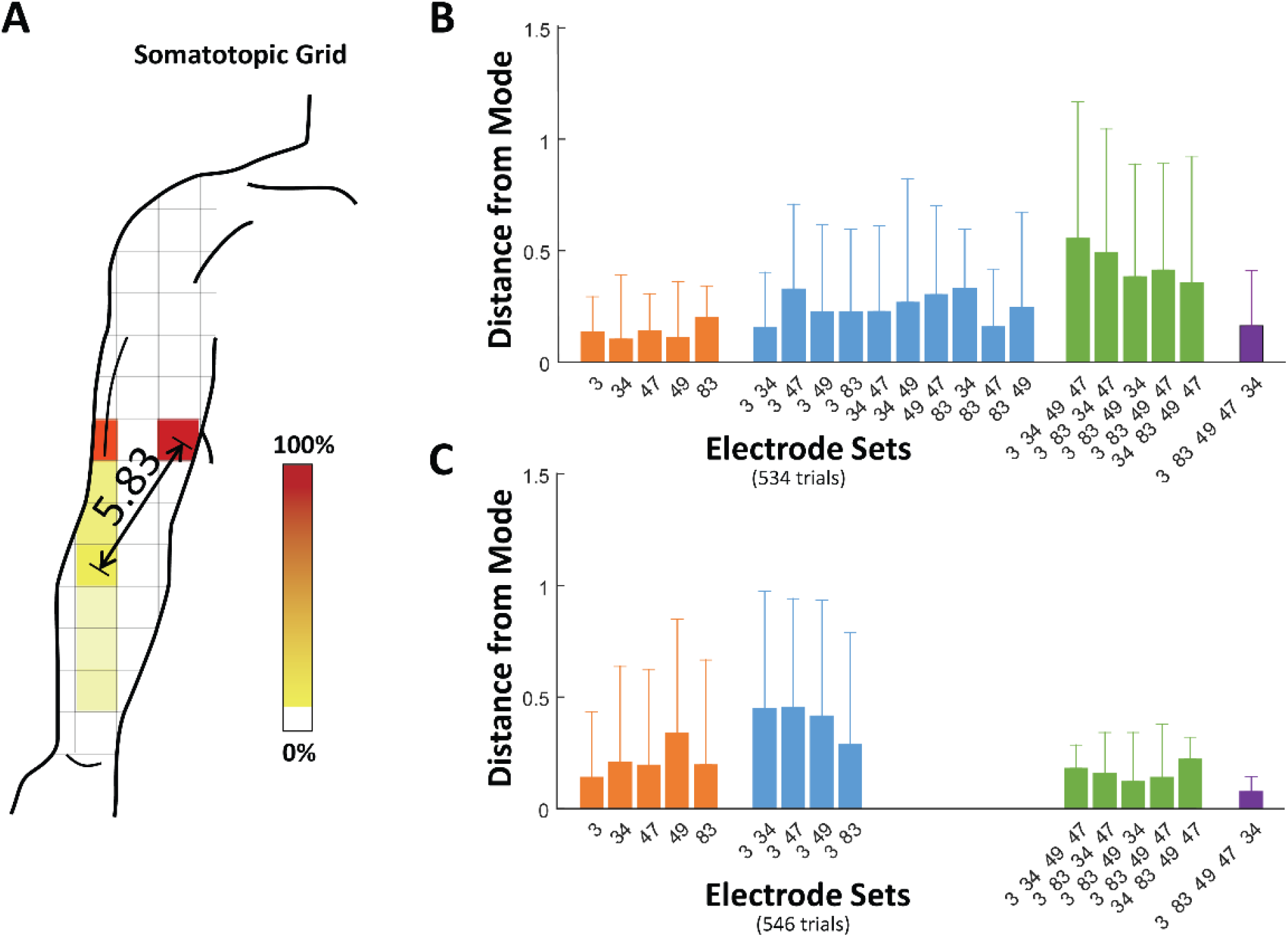
Variability of location of evoked sensations per electrode. The participant reported coordinates of the somatotopic location for each evoked sensation on a body map. Extremely low variance (mean values were below 0.6 for all cases) were calculated for somatotopic locations for all electrode combinations. **(A)** A illustrative example of reported locations from a single-channel stimulation pattern. The Euclidean grid distance was calculated between the most commonly reported location (the darkest color on the example body map) and all locations reported for a given electrode set. The colorbar shows the percentage of all reported sensations at each particular grid location. **(B)** Each colored bar illustrates the mean Euclidian grid distance and errors bars displaying one standard error of the mean. Each set of electrodes is shown individually, and the colors illustrate the number of electrodes in each set (one electrode – orange, two – blue, four – green, five – purple). Analysis is from data collected in Figure 4C. Only trials which elicited a sensation are included, thus the number of trials per bar varies between 9-57. **(C)** Identically analysis to Panel B is shown for data collected in Figure 4E. Due to limited experimental time, the first four sets of ten pairwise electrode combinations from Figure 4E were chosen. Only trials which elicited a sensation are included in this analysis, thus the number of trials varies between 24-53.

Using the verbal report task, each trial stimulated one ICMS pattern on a group of channel(s). If sensation was evoked, we asked the participant to report a description and somatotopic location (Figure 1A, bottom). All stimulation patterns were pseudo-randomly mixed trial by trial to control for possible habituation or hysteresis effects, and interleaved with catch trials (see methods).

### Charge density’s effect on detection thresholds

We analyzed the effects of charge density by measuring detection thresholds for each group of channels (Figure 4A) across a range of charge values (2nC to 20nC). Charge was calculated by integrating the positive amplitude of the ICMS pattern over the duration of the anodic phase and multiplying by the total number of pulses delivered. Our upper bound was determined by a charge safety limit of 20nC on a single electrode, in concert with the other stimulation parameters. We modulated charge by varying pulse-amplitudes (10-100µA) and the fixing pulse-width at 200us while stimulating for a one second duration at 300Hz.

Averaging over 10 trials per condition, we found the 70µA pulse-amplitude ICMS pattern cleared the 75% likelihood threshold for generating a sensation, for the average single-electrode in this group (Figure 4B). This 70µA pattern delivered 14nC of charge. For the multi-channel groups, we found a significant decrease in charge density required to meet the psychometric detection threshold (Figure 4D). Two-channel ICMS met this threshold at only 40µA (8nC per electrode), four-channel at 25µA (5nC), and five-channel at 20µA (5nC). Since a saturation effect was observed at 5nC per channel, this data suggests there exists both minimum amount of charge density required to evoke a sensation and a total charge requirement.

### Modulation of total charge delivered predicts detection thresholds

We analyzed the effects of total charge delivered by measuring detection thresholds for each group of channels across the same range of charge per electrode (2nC to 20nC). We modulated total charge by stimulating each type group using a range of pulse-widths (50ms-400ms) while fixing amplitude at 100µA for a one second duration at 300Hz.

On average, we found a 120µs pulse-width pattern met the psychometric detection threshold of 75% for single-channel ICMS (Figure 4C). This pattern delivered 12nC of total charge, comparable to 14nC value measured during the single-channel amplitude modulated patterns. For the four- and five-channel ICMS groups, we observed a significant decrease in required charge per electrode (Figure 4E). Both required only 8nC per electrode (a total charge of 32nC and 40nC, respectively) compared to 12nC per electrode for the single-channel ICMS pattern. This minimum was comparable to the 5nC per channel minimum required in the amplitude modulated dataset.

Unlike the pulse-amplitude modulation dataset, the psychometric detection curve for pulse-width modulation for the two-channel ICMS group did not significantly change compared to the single-channel group (Figure 4E, red line). Additionally, the total charge required to evoke a sensation during multi-channel ICMS was substantially lower for the pulse-amplitude modulation (16-25nC) than the pulse-width (32-40nC). While the psychometric curves for both pulse-amplitude and pulse-width paradigms suggested overlapping features (such as a minimum charge amount per channel), these differences suggested unique mechanisms of neural recruitment.

### Multi-channel ICMS patterns evoked more naturalistic sensations than single-channel

Previous studies have reported either only “natural” sensations [61] or a mixture of “natural” and “non-natural” sensations [12], [55] as described by human participants receiving single-channel stimulation patterns via ICMS or ECoG [62]. Without a clear explanation for these different results, it might be difficult to optimize stimulation patterns towards generating only “naturalistic” sensations for future sensory BMI applications. Understanding first principles of hypothesized stimulation effects may begin by correlating certain stimulation parameters with more “naturalistic” sensations. To test this hypothesis, we evaluated the elicitation frequency of “naturalistic” and “non-naturalistic” sensations with stimulation charge density (Figure 5).

From the datasets collected in Figures 4D, E, we categorized the participant’s reported descriptions of each evoked sensation as either “naturalistic” (e.g. touch, pinch, squeeze, etc) or “non-naturalistic” (e.g. “shock”). “Naturalistic” sensations were overwhelming reported for both single- and multi-channel ICMS patterns. However, a small but significant subset were classified as “non-natural” during single-channel ICMS in the pulse-amplitude modulated dataset (Figure 5A&B). Significance was tested between ratios of “natural” vs “non-natural” descriptions reported from single-vs multi-channel ICMS patterns (p<0.001, Wilcoxon rank sum test). Correlation between highly-focused charge ICMS delivery on a single-channel and an increase of “non-natural” sensations might suggest multi-channel ICMS may recruit cortical sensory networks more biomimetically.

### Stability in the reported location of evoked percepts

As multi-channel ICMS patterns likely activate a larger volume of cortex, the stability of a particular electrode’s evoked somatotopic location may be affected. Multi-channel stimulating electrodes were spaced 400µm apart. We found both single- and multi-channel ICMS patterns evoked sensations in highly stable somatotopic locations, while variability increased slightly during multi-channel stimulation. We quantified this variability from the dataset collected in Figure 4D,E.

For each channel (or group of channels), we plotted a heatmap of all reported locations on the body grid (Figure 6A). Using the grid coordinates of the body map (each grid space represented approximately 2×2 inch square on the body), we calculated the average Euclidian distance on the grid, between the most commonly reported grid location (shown in dark red) and all other reported locations (Figure 6A). By compiling a distribution of these distances, we calculated an estimate of the variability of reported body location for a given set of electrodes.

From the amplitude modulated dataset (Figure 4D), average grid distance values were extremely low (Figure 6B), trending between 0.25 and 0.50 as the number of electrodes increased. Similar results were found by analysis of pulse-width modulated data (Figure 6C). For all electrode groups, the average grid distance was less than 0.60, suggesting the robust stability of the evoked location between single and multi-channel paradigms.

Our participant did not report any locations of evoked percepts outside of the targeted body region (i.e. face, leg, torso) as we increased the number of electrodes. This result suggested cortical somatotopic representation of the arm is defuse enough for multiple neighboring microelectrodes to activate similar somatotopic regions. This could be due to large receptive fields or a lower density of innervation of the arm.

## DISCUSSION

Intra-cortical micro-stimulation (ICMS) has been proposed as an effective method for evoking naturalistic cutaneous and proprioceptive sensations for human patients suffering somatosensory loss. This study examined somatosensations evoked by multi-channel ICMS patterns, quantifying reaction times (RTs), verbal descriptions and somatotopic locations as reported by a tetraplegic human participant. We demonstrated RTs from naturalistic somatosensations evoked via multi-channel ICMS are comparable to naturally occurring vibrotactile sensations and significantly faster than visual stimulation (Figure 2). Across both implanted arrays, multi-channel ICMS patterns were more likely to evoke a sensation compared to single-channel ICMS patterns applied on the same electrodes (Figure 3). We found evidence of lossy integrative properties in cortical sensory networks, by comparing psychometric detection thresholds between single- and multi-channel ICMS (Figure 4). Utilizing spatial properties of multi-channel ICMS demonstrated that the charge density per electrode could be significantly reduced while simultaneously increasing the frequency of evoked “naturalistic” sensations (vs. “non-natural”) (Figure 5). Relatively stable localization of percepts of both single- and multi-channel ICMS were observed (Figure 6).

These results suggest BMI applications with highly temporal feedback requirements are possible (i.e. online, closed-loop, sensory feedback for dexterous object manipulation). Multi-channel ICMS patterns can reliably, precisely evoke stable naturalistic somatosensations with comparable cognitive latencies to naturally occurring somatosensations. Furthermore, the spatial organization of multi-channel ICMS patterns may be more biomimetic, leveraging the integrative properties of cortical networks and resulting a higher frequency of evoked “naturalistic” sensations.

### ICMS behaves similarly to natural sensory phenomena

Previous studies have yielded conflicting evidence whether sensations evoked via ICMS obey classic sensory psychophysical laws [12], [63], [64], such as whether the minimum perceptible change in stimulus intensity is proportional to the previous stimulus (Weber-Fechner Law [65]). Our results add evidence that ICMS evoked sensations do behave similarly to some psychophysical results from sensory intact human participants. Classic psychophysical experiments demonstrate decreased RTs with an increase of intensity of the stimulus [50], [66]– [70]. Our participant reported sensations with significantly higher intensities from four- and eight-channel ICMS patterns than sensations evoked from the single-channel ICMS. Correspondingly, RTs from those multi-channel ICMS patterns were significantly faster than RTs from the single-channel ICMS patterns.

### Reaction time task design

Classic reaction time experimental designs often cue the subject to press a button, flex their wrist, or make some other peripheral limb-motor intended action [71]. However, since our participant has full motor paralysis, peripheral limb motor actions are impossible. Saccadic eye movements to a fixed target are known to have a short delay to measured RTs during visual tasks [72]. However, previous studies have quantified this delay, measuring RTs with and without a saccade in response to a visual cue [72]. The measured range of the non-saccade RTs was between 217-303ms (95% confidence intervals), while upper limit of the saccade RTs range was substantially larger and longer (95% c.i.: 229-417ms). We do not see a correspondingly large range in our saccade RTs (c.i. 300-318ms), suggesting a minimal effect on our data.

Furthermore, each condition (visual, vibrotactile, ICMS) required the participant to saccade, so if present, this short delay is present in all conditions. Our work agrees with prior RTs measured in a non-human primate models for both single-[36], [42] and multi-channel ICMS [45]. Measured RTs for visual cues and vibrotactile stimuli in this study are similar to previously reported values for sensory intact humans pressing a button (210-400ms) [71], [73], [74].

### Reaction time measurements to electrical stimulation in human participants

Reaction times to direct cortical stimulation through both penetrating Utah micro-electrodes and ECoG electrodes have been measured in humans prior to our investigation. However, the comparison of RTs to ICMS and natural occurring sensory stimuli (visual or tactile) have been varied.

RTs measured from penetrating Utah micro-electrodes implanted chronically in a single participant with a spinal cord injury have been shown to be either comparable or slower than naturally occurring vibrotactile stimuli [46]. In this work, we show RTs from ICMS significantly faster than both visual and vibrotactile stimuli (Figure 2, four-channel ICMS). In comparison with previous work, this was achieved by stimulating on more electrodes simultaneously.

Electrocorticographic (ECoG) electrodes sit on the surface of the brain rather than in cortex where penetrating electrodes lay. Using a button press, researchers measured RTs to electrically evoked sensations approximately 100-200ms slower than response times to natural occurring stimuli [43]. The primary difference between our paradigm and previous studies are the type of electrode used. Implanted ECoG devices are much larger than the micro-electrode arrays used in this study and have a much lower electrode impedance (1-20kOhm). These properties affect both the current densities created by the stimulation pattern as well as the size, location and cell-types of the neural populations recruited. Both the lower impedance values and much large size of the ECoG electrodes requires miliamps of current to elicit a sensation, rather than microamps. The 3 orders of magnitude difference in charge, ensures that a much larger volume of cortical tissue is activated. ECoG electrode technology exists on completely different scale (5µm diameter exposed tip on a Utah array vs 2.5mm on a macro ECoG electrode) leading to much more defuse current spread across cortical networks.

Simultaneously stimulating a high number of channels allows us to achieve charge injection amounts that would not be safely possible on a single or pair of electrodes [56]–[58]. While this advantage increases the charge density on our micro-electrode arrays and likely recruits a larger pool of neurons, simply recruiting even larger neural populations (with another technology like ECoG) may not directly correspond to even faster reaction times than reported here.

### “Naturalistic” vs “non-naturalistic” sensations

Previous stimulation studies have described either only “natural” sensations [3] or a mixture of “natural” and “non-natural” sensations [12], [55] reported by human participants receiving single-channel stimulation patterns both in ICMS and ECoG [62]. “Non-natural” sensations have been reported both immediately post implantation and throughout the study periods [12], [35].

Several plausible hypotheses have been suggested: initial tissue response due to implantation of the array, degradation of tissue-electrode interface, tissue scaring around the electrodes over time, localization of the array in cortex, and non-natural recruitment of neural populations from stimulation patterns. It is beyond the scope of this study to validate or reject these hypotheses; however, our data shows evidence that multi-channel stimulation patterns preferentially evoke “natural” sensations (Figure 5). The underlying variable here, charge density, could be linked to a more natural recruitment of neural populations. Rather than bombarding a very tiny volume of tissue with free electrons, spreading the same amount of charge across a volume double or quadruple in size may activate the neural population in a more biomimetic manner.

## MATERIALS AND METHODS

### Participant

A human participant with C5 level spinal cord injury was consented and chronically implanted with Utah arrays [47], [48]. Two SIROF tipped 48-channel Utah arrays [75] were placed in primary sensory cortex (S1), targeting the hand and arm region [3]. Given the high level of injury, the participant had limited motor use of the shoulders and bicep muscles, with little or no volitional control below this level of their body. Cutaneous and proprioceptive sensory perception was also severely comprised; however, localized regions of the elbow, right thumb and inner right arm had intact sensation. Further details about the participant, implantation procedure, surgical planning, implant locations and methods can be found in [3]. All human subject research, experimental design and biomedical devices used in this study were approved by the Institutional Review Boards (IRB) of the California Institute of Technology, University of Southern California, and Rancho Los Amigos National Rehabilitation Hospital for collection of this data.

### Reaction time task

Using a simple reaction time (RT) paradigm [66]–[68], we instructed the participant to respond as quickly as possible after the onset of several different stimuli types. We compare RTs between visual stimuli, vibrotactile stimuli and a variety of intra-cortical micro-stimulation (ICMS) encoding paradigms to illustrate the cognitive processing times required for each. Since the participant has limited volitional control of their arms and hands, we used a commercially available eye-tracker (Tobii 4C - Tobii Gaming, Inc) to measure reaction time performance.

The task had four phases: an inter-trial interval, cue, fixation, and stimulus phase (Figure 1A). After the ITI phase, the participant was cued to the type of stimulus to be delivered during the upcoming trial. Thus no additional cognitive latency would be introduced from attending to three possible sources of stimuli.

During the fixation phase, two targets were present, a fixation circle at the center of the screen and a saccade target to the right. The participant was instructed to fixate on the center circle on a video monitor (120FPS refresh rate) and saccade rightward, immediately after the onset of the stimulus presentation. This fixation was enforced for a variable inter-trial-interval of 1.5-3 seconds. If fixation was broken, this timer would reset.

After the fixation period, the participant experienced one of three types of stimuli: visual, vibrotactile or an ICMS pattern (Figure 1A). Trials with each stimulus type were pseudo-randomly interleaved and balanced such the frequency presentation of each stimulus type was equal during a block of trials.

The visual stimulus was a green circle, immediately replacing the blue fixation circle. During the vibrotactile condition, the fixation circle remained unchanged and the participant saccaded after experiencing an evoked natural sensation via the vibrotactile motor (C3 Tactor – Engineering Acoustics, Inc). The location of the vibrotactile motor was placed close to the approximate location of the evoked ICMS sensation on the arm. Details regarding placement can be found below.

During the ICMS conditions for the reaction time task, the participant responded to an evoked artificial sensation. We measured the participant’s reaction time to three different ICMS patterns. All patterns applied a 200ms second duration pattern, 200µsec pulse-width, 300Hz stimulation frequency pattern to the electrodes selected. The first ICMS pattern applied this pattern with a 100µA pulse-amplitude to a single electrode, and 2nd and 3rd patterns applied this same pattern to a group of 4 and 8 neighboring electrodes respectively. Electrical safety constraints [56]–[58] on the amount of instantaneous charge injected and the instantaneous charge density limited our 8 channel stimulation pattern to 90µA per channel.

### ICMS pattern generation

Intra-cortical micro-stimulation (ICMS) patterns were generated using four key parameters [59]: pulse amplitude, pulse-width, stimulation frequency and total stimulation duration (Figure 1C). Pulse-amplitude ranged from 1-100µA, pulse-width from 44-400µsec, while the stimulation frequency was fixed at 300Hz and duration lasted up to 1s. A 53µsec inter-phase interval was placed between the leading cathodic and preceding anodic phase of each stimulation pulse. Each pulse was charge balanced for safety, with symmetrical cathodic and anodic phases.

Electrical charge (Q) is a measure of current injected over a given time period. In this paper, charge is measured in nanoCoulombs (nC) and is calculated as the pulse-amplitude (µA) * cathodic pulse-width (µs) * number of delivered pulses * number of electrodes. The number of delivered pulses is equal to the stimulation frequency (Hz) * stimulation duration (s).

### Selection of electrodes

Groups of electrodes were selected for the reaction time experiments (Figure 1D) and single-vs multi-channel ICMS patterns (Figure 4A). Prior to and during the course of the experimental timeline of this paper, we aggregated data from many different somatosensory stimulation experiments in our lab to generate a heatmap of the probability of evoking a sensation on any electrode on both our arrays (Figure 1B). This heatmap combines data across a variety of stimulation patterns, stimulus presentations and task designs while also controlling for an equal number of stimulus presentations per electrode. For this aggregate heatmap, we have 32±2 (mean, standard deviation) trials per electrode. For the experiments in this paper, we used this data to select electrodes groups with a high probability of evoking a sensation.

### Placement of vibrotactile sensation

A somatotopic sensitivity map of the arm and hand region was probed using Semmes and Weinstein Monofilaments (SWM) [76] while the participant wore a blindfold. These filaments measure sensitivity to touch by applying a precise amount of force (0.07, 0.4, 2, 4, and 300 grams, manufactured with <5% deviation). These levels correspond to Normal, Diminished Light Touch, Diminished Protective Sensation, Loss of Protective Sensation and Deep Pressure Sensation Only [77]. Any region which the participant had no response to any level of force were considered insensate. Reported somatotopic locations on the forearm and hand typically evoked by our ICMS patterns were evaluated with the full range of SWMs. The vibrotactile sensor was placed on two locations sensitive in the Normal range (Figure 2A, black “X” and right, mid cheek).

### Verbal report task

This behavioral task [3] measured the qualitative aspects of each evoked sensation and captured the participants perceived description and somatotopic location (Figure 3A). Each trial was separated by a short inter-trial interval (2 secs) followed by an ICMS stimulus presentation. A visual cue (a green fixation circle at the center of screen) illuminated while ICMS was given. After each presentation, the participant verbally responded if a sensations was perceived. If yes, a qualitative description (i.e tap, touch, pinch, squeeze, etc) and a somatotopic location (grid coordinates from the arm/hand map) were reported. See Figure 3 for an example.

### Catch trials

We used catch trials to confirm the participant was using the stimulation and no other stimuli to determine the perceived sensations. Visual input remained the same during the task while the stimulator delivered no current during these trials. Catch trials were pseudo-randomly interleaved at an approximate 5% rate. We collected approximately 3500 trials in over 50 blocks, with 175 catch trials and zero false positives. The only observance of false positives occurred when the participant was briefly on a medication with a side effect of severe lethargy. It was readily apparent the participant was unable to complete the task, and data collected on these days was not included in the analysis.

### Hardware and data collection

A Blackrock Microsystems Inc, Neural Biopotiental Signal Processor (NSP) captured relevant neural data at 30kHz sampling rate and the CereStim96 device generated micro-stimulation patterns delivered to the S1 arrays. A custom computer program (MATLAB) utilized PyschToolbox to display visual cues during the task, log relevant behavioral data and participant responses, and program the Blackrock CereStim96 with each trial’s intended stimulation pattern. Tobii eye-tracking data was sampled at 66Hz, aligned with stimulus presentation and recorded in the Matlab program.

### Statistical analysis

Reaction time was measured as the time from the onset of a stimulus presentation to the beginning of a detected saccade. Initiation of a saccade was measured using an offline analysis of eye-tracking trajectories during the stimuli presentation. A boundary around the fixation circle was drawn and the time point was measured at which the eye’s rightward trajectory crossed this line. The participant was given a practice session each day that data was collected such that values measured would reflect peak performance.

Reaction time distributions were tested for significance by performing a pairwise Wilcoxon rank-sum hypothesis test for two samples drawn from independent continuous distributions [78]. This test does not assume any parametric conditions on the two distributions and is robust to differing sample sizes. We corrected for multiple comparisons with the Bonferrioni correction method [79].

## Data Availability

All data produced in the present study are available upon reasonable request to the authors

## Supplementary Materials

None.

## Acknowledgments

The authors would like to acknowledge FG for his efforts and engagement in the clinical study, the clinical staff at Rancho Los Amigos and the technicians at Pierce Congregate Living and for their work and dedication during the experimental sessions.

## Funding

Boswell Foundation

T&C Chen Brain-machine Interface Center

NIH/NINDS Grant U01NS098975

NIH/NINDS Grant U01NS123127

Craig F. Neilson Foundation

## Author contributions

Conceptualization: DB, LB, CYL, RAA

Methodology: DB, LB, BL, CYL, RAA

Investigation: DB, LB

Visualization: DB

Funding acquisition: DB, LB, CYL, RAA

Resources: DB, LB, KP, BL, CYL, RAA

Project administration: KP, RAA

Supervision: RAA

Writing – original draft: DB

Writing – review & editing: DB, RAA

## Data and materials availability

All data used to generate figures are available upon reasonable request.

